# Differentiated care for youth across the HIV care cascade in Zimbabwe

**DOI:** 10.1101/2023.10.11.23296905

**Authors:** Chido Dziva Chikwari, Katharina Kranzer, Victoria Simms, Amani Patel, Mandikudza Tembo, Owen Mugurungi, Edwin Sibanda, Onismo Mufare, Lilian Ndlovu, Joice Muzangwa, Rumbidzayi Vundla, Abigail Chibaya, Richard Hayes, Constance Mackworth-Young, Sarah Bernays, Constancia Mavodza, Fadzanayi Hove, Tsitsi Bandason, Ethel Dauya, Rashida Abbas Ferrand

## Abstract

**Introduction:** Youth living with HIV are at higher risk than adults of disengaging from HIV care. Differentiated models of care such as community antiretroviral therapy (ART) may improve treatment outcomes, including viral suppression.

**Methods:** This study was nested in a cluster-randomised controlled trial (CHIEDZA: Clinicaltrials.gov, Registration Number: NCT03719521) which was conducted in Harare, Bulawayo and Mashonaland East Provinces in Zimbabwe and aimed to investigate the impact of a youth-friendly community-based package of HIV services, integrated with sexual and reproductive health services for youth (16–24 years), on population-level HIV viral load (VL). HIV services included HIV testing, ART initiation and continuous care, VL testing, and adherence support. Coverage percentages across the HIV care cascade were analysed.

**Results:** Overall 377 youth were newly diagnosed with HIV at CHIEDZA, linkage to HIV care was confirmed for 265 (70.7%, 234 accessed care at CHIEDZA and 31 with other providers); 250 (94.3%) started ART. Among those starting ART at CHIEDZA attrition within 6 months of starting ART was 38% and viral suppression (<1000 copies/ml) among those who had a test at 6 months was 90%. In addition 1162 youth already diagnosed with HIV accessed CHIEDZA; 714 (61.4%) had a VL test, of those 565 (79.1%) were virally suppressed.

**Conclusion:** This study shows that provision of differentiated services for youth in the community is feasible. Linkage to care and retention during the initial months of ART was the main challenge as has been shown in other studies. Retention throughout the HIV-care journey for youth needs to become a priority programme goal to achieve the ambitious 95-95-95 UNAIDS targets.

## Introduction

In 2019, an estimated 3.9 million young people (aged 15-24 years) were living with HIV globally, the majority in Southern Africa.(1) Young people continue to be the age group with the highest HIV incidence, especially among women who experience three times higher incidence than males of the same age.(2)

In 2014, UNAIDS set the 90−90−90 target aiming for 90% of people living with HIV to be aware of their HIV status, 90% of people diagnosed with HIV to be on antiretroviral therapy (ART), and 90% of those people on ART to have suppressed HIV viral load by 2020.(^3^) These targets were followed by the even more ambitious 95-95-95 targets, to be achieved by 2025.(4) While Southern African countries have made great progress towards the 90-90-90 targets, progress has been much slower for young people living with HIV in the region.(5–10) Young people face challenges across each step of the HIV care cascade beginning with knowing their HIV status through testing, timely and continuous access to treatment, and optimal care outcomes including viral suppression.(11–16) Successful interventions to increase testing and knowledge of HIV status among young people have included community-based HIV testing services including HIV self-testing offered at home, through outreach programmes, at educational facilities and at fixed community locations.(17–21) Few studies aimed at addressing the gap for young people in the ‘first 95’ (knowledge of HIV status) have also reported linkage to and continuous engagement in HIV care.(17)

Although retention in long-term HIV care remains one of the most difficult challenges across all age groups, young people are more likely than any other age group to disengage. Barriers to their retention include financial dependency, stigma, health services that are not youth friendly and incompatible with school attendance, and adolescent development adjustments as they transition to adulthood.(22–24) Importantly the bulk of attrition occurs before ART initiation and within the first 6 months on treatment.(12, 25) There is a recognised need for concentrated attention on health-care approaches to HIV service delivery for young people to bridge these gaps.(26)

Simplified and patient-centred care options tailored to each stage of the patient care journey (‘differentiated models of service delivery’) aimed at improving engagement and retention in care have been developed for and tested among adults living with HIV. However, few differentiated models of care specifically targeting young people have been described and data on the implementation and effectiveness of these interventions remain scarce.(27)

In this paper, we describe the results of a differentiated model of service delivery offering community-based HIV and VL testing, linkage to care, ART initiation, HIV care and health-care worker managed adherence groups for young people living with HIV aged 16-24 years in three provinces in Zimbabwe.

## Methods

### Trial design, intervention and setting

This study was nested within a cluster randomised trial (CHEIDZA) conducted in three urban and peri urban provinces across Zimbabwe, each province containing 8 clusters randomised 4:4 to control (existing health services) or intervention clusters.(28) The trial aimed to investigate the impact of a comprehensive community-based package of HIV services, integrated with sexual and reproductive health services and general health counselling for youth aged 16–24 years, on population-level HIV viral suppression. A cluster was defined as a geographically demarcated area with a multi-purpose community centre. In intervention clusters, weekly integrated HIV and sexual and reproductive health services were delivered from this centre to cluster residents aged 16-24 years. Services included HIV testing, HIV treatment and adherence support for those living with HIV, contraception, menstrual hygiene management, syndromic management of sexually transmitted infections, risk reduction and general health counselling offered by a multidisciplinary team of service providers over 30 months.(28)

The intervention was specifically configured to be “youth friendly” i.e. able to effectively attract youth, meet their needs responsively and retain them in care. Part of the youth friendliness was a focus on intervention providers; the intervention team was selected based on prior experience of working in communities and with youth. A training programme on each of the intervention components was combined with training on provision of youth friendly services, particularly focusing on communication and counselling that is appropriate to age and maturity, LGBTQI+ sensitivity and attitudinal training specifically emphasizing respect, confidentiality, non-judgement, and relatability. Debrief meetings for the intervention teams were held every 1-2 weeks and incorporated problem-solving, discussion of complex cases and operational issues to ensure that intervention providers were supervised and mentored. The start of the intervention period was staggered, with Harare province starting on 1 April 2019, followed three and six months later in Bulawayo and Mashonaland East provinces respectively. The intervention period ended on 31 March 2022. All services were voluntary and offered free of cost.

### HIV testing and care services

HIV testing was conducted according to national guidelines.(29) Those who tested HIV-positive were offered a choice of being referred to a primary care clinic (PCC) of their choice (accompanied to the PCC by a CHIEDZA member of staff to help facilitate linkage to care) or of accessing care in the community through CHIEDZA. If the latter was selected, the young person was assigned a national programme number and their HIV records were maintained at the PCC. For those who accessed care through CHIEDZA ART was supplied by the PCC through the national HIV programme. CHIEDZA staff updated the PCC data when they collected ART for supply through CHIEDZA. This ensured that young people remained part of the national HIV programme allowing them to transition to receiving care from any PCC of their choice at the end of the trial. HIV treatment was provided according to national guidelines(30) and there were referral pathways to a health facility for any clinical indications (e.g., severe toxicity, incident symptoms, suspected treatment failure). A point-of-care CD4 count was done at time of diagnosis or ART initiation at the CHIEDZA site. Those with a CD4 count <100 cells/uL had a serum cryptococcal antigen test. All young people initiated on ART had a WHO tuberculosis symptom screen. Those with a positive symptom screen were further investigated for tuberculosis as clinically indicated including sputum Xpert MTB/Rif testing, chest X-rays and/or investigations for extrapulmonary tuberculosis done offsite at a local laboratory.

Young people newly diagnosed with HIV or known positive but not on ART were followed by telephone and / or home visits if they did not attend scheduled appointments. There was no defined protocol for follow up, however, multiple attempts were made to contact a person and facilitate linkage to care.

Young people living with HIV and already accessing care outside CHIEDZA were encouraged to remain with their routine care provider. However, those who wished to transfer care to CHIEDZA were able to do so.

Young people living with HIV (regardless of whether they accessed HIV care through CHIEDZA or not) were offered free HIV VL testing after 6 months on ART (from September 2019). VL testing was done offsite with results available within approximately one week. Enhanced adherence counselling was provided at CHIEDZA if not virally supressed (>1000 copies/ml) as per Zimbabwe national guidelines. All youth living with HIV were invited to join CHIEDZA Adolescent Peer Support (CAPS) group that were modelled on the existing Community ART Refill Groups (CARGs) implemented in Zimbabwe .(31) CAPS comprised a group of young people with HIV who met on a regular basis to discuss issues relevant to youth and engage in social activities as a model of receiving peer support. The group facilitator was a CHIEDZA staff member.

### Outcomes

Clients were defined as previously diagnosed with HIV based on self-report, and as newly diagnosed if they self-reported negative or unknown HIV status and then tested HIV positive at the CHIEDZA service. Clients who did not link to care at CHIEDZA were asked to leave a contact phone number, and were followed up to determine whether they linked to care elsewhere. Clients were coded as linked to care elsewhere if they ever reported that they were receiving HIV service from another provider, with no time limit. Similarly, clients were coded as linked to care at CHIEDZA if they ever took up HIV care at CHIEDZA. Clients who did not leave contact information were coded as having unknown outcome and those who were contacted at least once but stopped responding thereafter were coded as lost to follow-up. Clients living with HIV, whether newly diagnosed or previously diagnosed, who opted to receive HIV care at CHIEDZA were allocated a 4-digit ID number. They were referred to as the CHIEDZA HIV cohort.

All clients on ART, including those not linked to care at CHIEDZA, were eligible for a VL test after 6 months on ART, however, clients in the CHIEDZA HIV cohort were ineligible for VL testing if they transferred elsewhere before completing 6 months on ART at CHIEDZA, or if the CHIEDZA service closed before they completed 6 months on ART. In addition, clients who received HIV care elsewhere were not eligible for VL testing if they attended CHIEDZA before September 2019. Viral suppression was defined as a VL <1000 copies/ml.

### Data management and statistical analysis

Information on CHIEDZA service uptake and minimal demographic information, (sex, date of birth) was collected from all CHIEDZA attendees via an electronic tablet and linked to a fingerprint ID. HIV cohort members were asked to complete an additional electronic case report form (CRF) at baseline and at each subsequent visit capturing sociodemographic and clinical information. Separate paper CRFs captured VL and CD4 test results. These were entered into a purpose-built database by data clerks.

Clients in the CHIEDZA HIV cohort were identified by their 4-digit ID number. Those who received HIV care elsewhere were identified by the first eight characters of their fingerprint ID.

The CHIEDZA staff followed up clients to ascertain whether they had linked to care and initiated ART elsewhere, or whether they had relocated, refused to respond, were not ready to initiate ART, or had been referred for another condition. Participants who could not be traced because they did not supply contact details were coded unknown. This information was compiled in a spreadsheet by the data manager and merged with the main dataset.

### Ethics

Ethical approval for the CHIEDZA study was obtained from the Medical Research Council of Zimbabwe [MRCZ/A/2387], the Biomedical Research and Training Institute Institutional Review Board [AP149/2018] and the London School of Hygiene & Tropical Medicine Ethics Committee [16124/RR/11602]. All intervention attendees provided verbal consent for services. The requirement for guardian consent to access services for 16–18-year-olds was waived by the ethics committees.

## Results

A total of 377 young people were newly diagnosed with HIV at CHIEDZA, while an additional 1162 young people living with HIV accessed the CHIEDZA service during the intervention (April 2019-May 2022) (Figure 1). Among those newly diagnosed 265/377 (70.3%) decided to access care at CHIEDZA (n=234) or with other service providers (n=31). Among the remaining 112 (29.7%) 74 did not link to care and for 38 linkage was unknown (Figure 1, Table 1). Young people who did not link to care: i) said they were not ready to start ART (n=11), ii) refused to be contacted again, but were not on ART at the last contact (n=14) or iii) were initially contactable following the positive HIV test, but were eventually lost to follow-up after multiple contacts encouraging them to link to care (n=49). Those whose linkage status was unknown: i) had relocated to another community (n=4), ii) lived outside the intervention cluster (n=5) iii) had been referred to other service providers (n=8) or iv) had left the CHIEDZA service after having tested HIV positive without providing any contact information (n=21).

**Figure 1:**
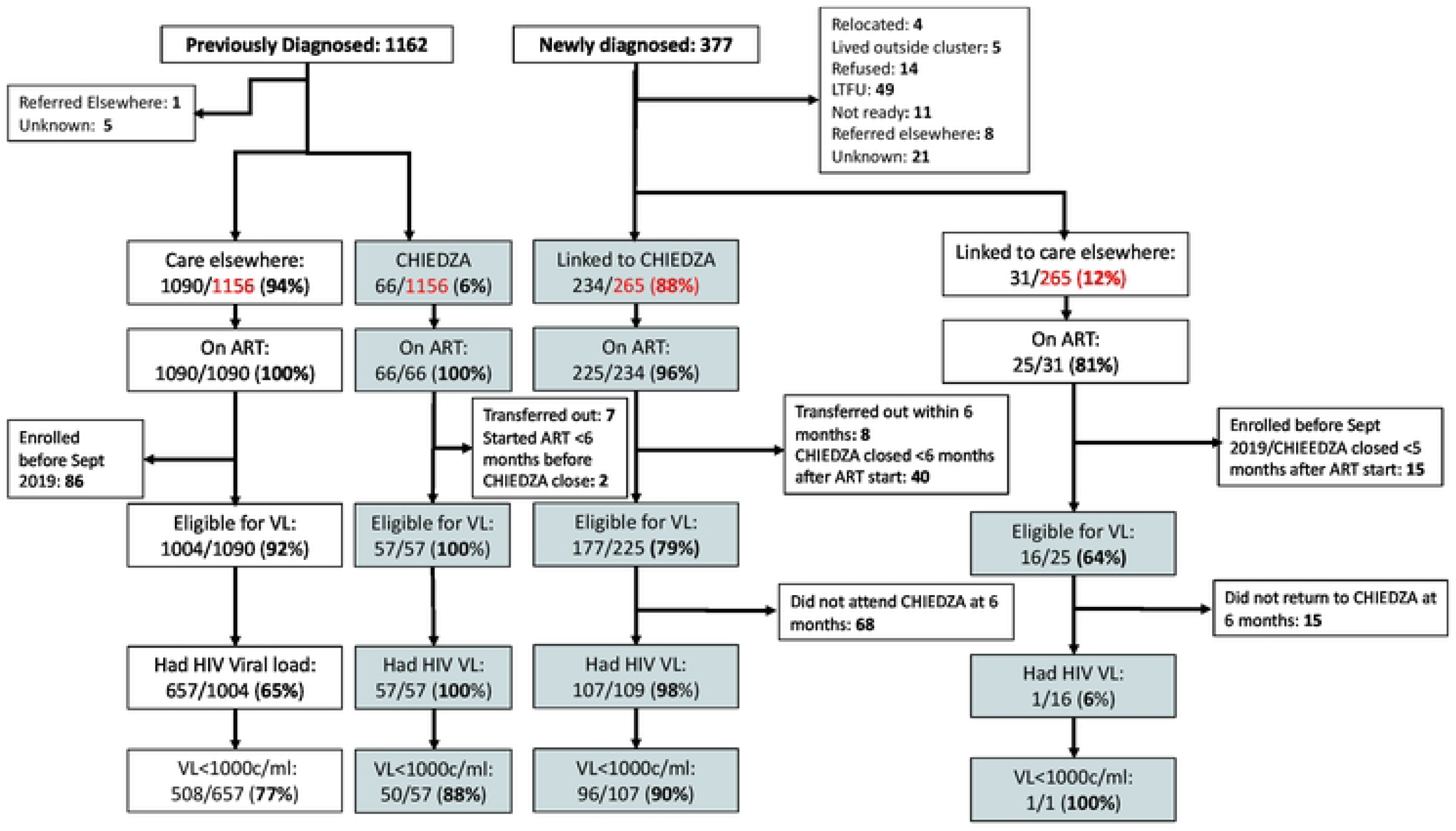
Flowchart of young people livingwith HIV who accessed CHIEDZA services

**Table 1:** Baseline characteristics of those newly diagnosed and previously diagnosed with HIV.

| Characteristics |  | Newly diagnosed, n (row %) |  |  |  |  | Previously diagnosed, n (row %) |  |  |
| --- | --- | --- | --- | --- | --- | --- | --- | --- | --- |
|  |  | Total (n=377) | Linkage unknown (n=38) | No linkage (n=74) | Linked elsewhere (n=31) | Linked to CHIEDZA (n=234) | Total (n=1156)* | Transferred care to CHIEDZA (n=66) | In care elsewhere (n=1090) |
| Age (years) | 16-19 | 114 | 17 (14.9) | 31 (27.2) | 7 (6.1) | 59 (51.8) | 328 | 22 (6.7) | 306 (93.3) |
|  | 20-24 | 263 | 21 (8.0) | 43 (16.4) | 24 (9.1) | 175 (66.5) | 828 | 44 (5.3) | 784 (94.7) |
| Sex | Male | 41 | 8 (19.5) | 8 (19.5) | 1 (2.4) | 24 (58.5) | 121 | 3 (2.5) | 118 (97.5) |
|  | Female | 336 | 30 (8.9) | 66 (19.6) | 30 (8.9) | 210 (62.5) | 1035 | 63 (6.1) | 972 (93.9) |
| Province | Harare | 124 | 12 (9.7) | 19 (15.3) | 12 (9.7) | 81 (65.3) | 365 | 10 (2.7) | 355 (97.3) |
|  | Bulawayo | 92 | 6 (6.5) | 29 (31.5) | 3 (3.3) | 54 (58.7) | 353 | 3 (0.9) | 350 (98.3) |
|  | Mashonaland East | 161 | 20 (12.4) | 26 (16.2) | 16 (9.9) | 99 (61.5) | 438 | 53 (12.1) | 385 (87.5) |
| Self-reported HIV status | Unknown | 163 | 23 (14.1) | 28 (17.2) | 13 (8.0) | 99 (60.7) | NA | NA | NA |
|  | Tested HIV-ve >6mths ago | 173 | 12 (6.9) | 36 (20.8) | 16 (9.3) | 109 (63.0) | NA | NA | NA |
|  | Tested HIV-ve <6mths ago | 41 | 3 (7.3) | 10 (24.4) | 2 (4.9) | 26 (63.4) | NA | NA | NA |

\*Excluding 6 for whom ART treatment status is unknown

A minority of young people newly diagnosed with HIV and linked to care (9/234 (3.8%) linking to CHIEDZA and 6/31 (19.4%) linked to other providers) did not start ART during follow-up. Among those who started ART in CHIEDZA, 48 were either transferred out or did not have enough follow-up time (<6 months) to be eligible for a VL test. In addition 68/177 (38%) did not attend follow-up visits at CHIEDZA six months after starting ART and their ART status thereafter was unknown. Among those who stayed in care with CHIEDZA >6 months after starting ART VL suppression was 90% (96/107). Using <20 copies per ml as the cut off; viral suppression in this group was 78% (83/107).

A small number (66/1162, 5.7%) of young people previously diagnosed with HIV transferred their care to CHIEDZA, their viral suppression was 88% (50/57). Among eligible young people previously diagnosed with HIV and receiving care from other providers 65.4%) (657/1004) were offered and agreed to have a viral load test. Viral suppression was 77% (508/657).

Table 1 describes the baseline characteristics of young people newly diagnosed with HIV and those with a known HIV positive status. The majority of young people identified with HIV were women (newly diagnosed 336/377, 89% and previously diagnosed 1035/1156, 89%) and the majority were aged 20-24 (newly diagnosed 263/377, 70%, and previously diagnosed 828/1156, 76%). 16-19 year olds newly diagnosed with HIV were less likely to link to care than those aged ≥20 years. CD4 counts were available for 228/234 newly diagnosed young people linked to care in CHIEDZA: 22 (20%), 101 (44%) and 105 (46%) had CD4 counts of <200, 200-499, >500 cells/uL respectively. Of the 4 young people with a CD4 count <100 cells/uL 3 had a serum cryptococcal antigen test, and all of those tests were negative. No young person was diagnosed with tuberculosis through CHIEDZA.

Few young people already in care with other providers transferred their care to CHIEDZA. Transfer was more frequent in Mashonaland East 53/428 (12%) compared to other provinces. Of 1539 young people living with HIV who accessed CHIEDZA 323 (21%) registered with CAPS. CAPs registration was more frequent among those accessing HIV care at CHIEDZA (110/300, 37%) compared to those accessing HIV care from other providers (213/111, 19%). A total of 32 CAPS sessions with a total of 734 attendances were held across the three provinces (Figure 2). National and local lockdowns due to the COVID-19 pandemic severely affected CAPS groups both with regards to hosting the events and attendance.

**Figure 2:**
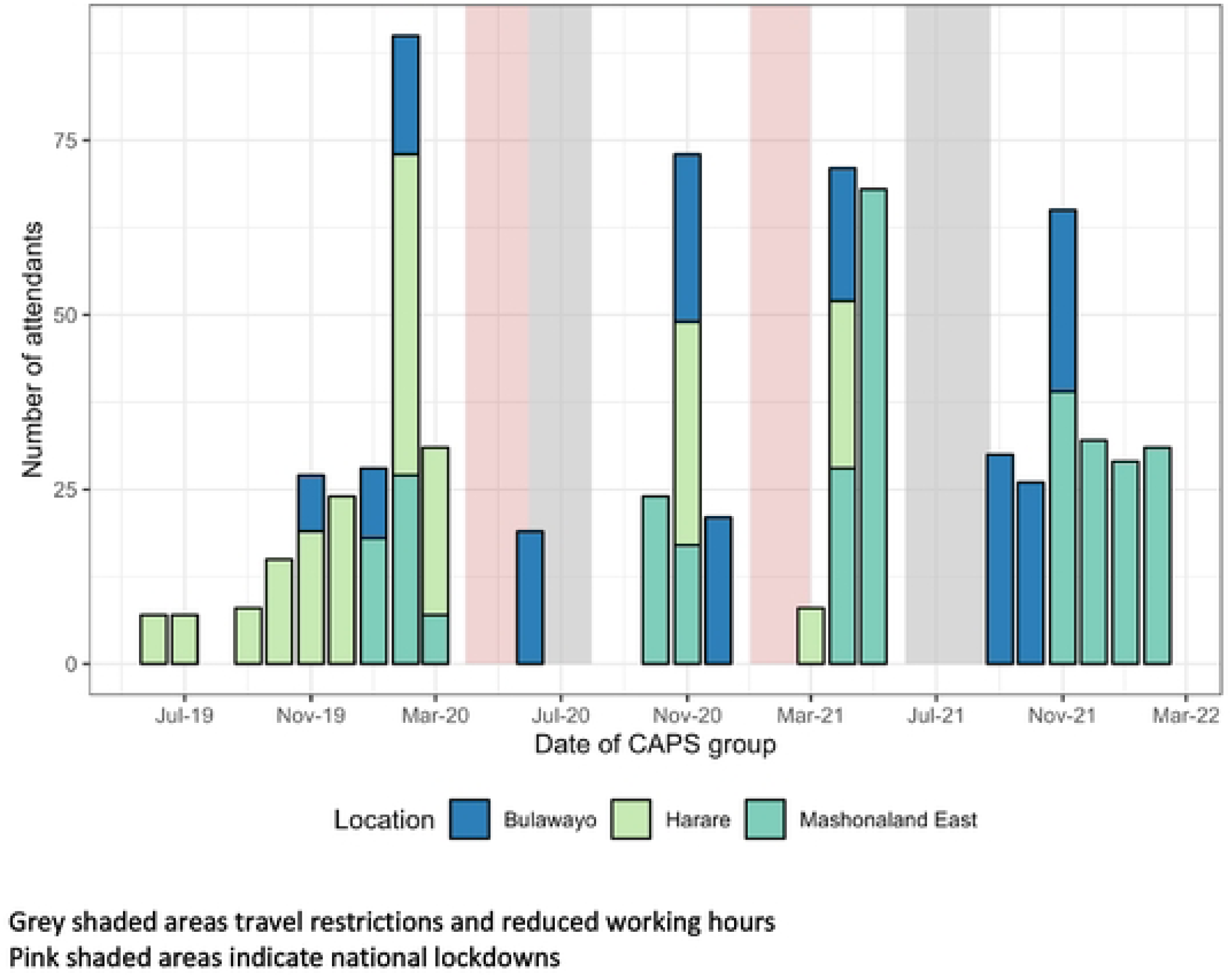
Number and timing of CHIEDZA young people peer support (CAPS) groups

## Discussion

We report outcomes across the HIV care cascade from a differentiated service delivery model for young people and co-developed with young people. The CHIEDZA intervention offered HIV testing integrated with sexual and reproductive health services, ART initiation, treatment monitoring (HIV VL testing) and adherence support (CAPS groups) in the community. Importantly the service was not just youth friendly, but offered choice and service providers respected young people’s autonomy. For example young people newly diagnosed with HIV who linked to care within CHIEDZA were provided with continuous care and services regardless of their decision on whether to start ART. Among young people who were newly diagnosed with HIV, 70.3% were confirmed to have linked to care, among those linked to care 94.3% started ART and among those starting ART at CHIEDZA and who remained in care beyond six months 89.7% were virally suppressed. However, a considerable proportion of young people either did not link to care or their linkage status was unknown. Some of them found it difficult to accept the diagnosis and either refused further contact or left the services without providing any contact information. Importantly 38% of young people newly diagnosed with HIV disengaged from care within the first six months of having initiated ART. In addition early attrition from ART is common among young people and has been reported from other countries and settings.(11, 12, 25, 32) Part of investing in retaining youth newly diagnosed with HIV in care is potentially doing further work in preparing young people for the implications of testing by ensuring there is more in place at the point of testing to improve HIV treatment literacy and protect against individuals disengaging after testing.(11) This could be nested within HIV pre-test counselling and incorporate messaging about viral suppression and U=U.

Linkage to care following HIV testing in the community is challenging. Those accessing community HIV testing services are less drawn to clinic-based services otherwise they would have tested at a clinic. Additionally, community-based services often cannot provide the same level of services with regards to frequency (5-6 days per week) and working hours as clinic-based services and therefore linkage to care and ART initiation is rarely offered as part of community-based services. A systematic review including 14 studies focused on community-based HIV testing reported proportions linked-to-care ranging from 10–79% over 1–12 months of observation.(33) None of the studies included in the review focused exclusively on young people and adolescents (16-17 year olds) were generally excluded. A more recent study “PopART for Youth” (P-ART-Y) delivered a combination HIV prevention package to 10-19 year olds in Zambia and South Africa via a door-to-door approach. The prevention package included HIV testing, supported linkage to care for those living with HIV and ongoing ART adherence support. In a before/after comparison ART coverage among adolescents living with HIV increased from 61.3% in Zambia and 65.6% in South Africa to 78.7% in Zambia and 87.8% in South Africa after one year of the intervention. While the intervention had the biggest impact on the first 90% (adolescents with HIV knowing their status), linkage to care must have been effective in order to increase ART coverage overall.(17)

Few studies have investigated the feasibility and outcomes of ART initiation in the community and those did focused on adults only.(34–36) Two studies investigated the feasibility of initiating ART in the community either following an HIV self-test in Malawi(35) or in the context of door-to-door HIV testing in Lesotho(36) with follow-up care provided at the clinic. ART initiation was increased in both studies and viral suppression (<100 copies per ml) 12 months post diagnosis was higher (50.4%) in the community ART initiation group compared to the standard of care group (34.3%) in Lesotho. Barnabas et al investigated the effect of community-compared to clinic-based ART delivery (i.e. initiation and ongoing care) on viral suppression among adults newly diagnosed with HIV in Uganda and South Africa.(34) Participants enrolled in the trial were diagnosed at clinics, through HIV testing at community locations and at home including distribution of HIV self-test kits. Data on linkage to care were not available. However among those initiating ART viral suppression (<20 copies per ml) at 12 months was 74% among those receiving community-based ART compared to 63% among those receiving clinic-based ART. Reanalysing our data using the same cut-point for viral suppression (<20 copies per ml) and among those who did have an HIV viral load the prevalence of viral suppression in young people newly diagnosed with HIV was 78% (83/107). This is comparable with the results achieved in adults in Uganda and South Africa, however, it does not take into consideration young people who have not been linked to care.

The CHIEDZA trial actively sought to limit the disruption of existing service provision. Thus young people previously diagnosed with HIV and in care with other service providers were encouraged to remain with their providers. This was largely achieved as evidenced by only 6% of young people living with HIV transferring their care to CHIEDZA. However, CHIEDZA offered additional services for those known to be HIV positive including community based HIV VL testing, adherence counselling if found to be unsuppressed and adherence and peer support through the CAPS groups. HIV VL testing at CHIEDZA was taken up by two thirds of eligible young people known to be HIV positive and 77.3% were found to be virologically suppressed which is in line with most of the estimates in this age group.(14, 37) A minority of young people newly diagnosed or known HIV positive registered with CAPS. Possible reasons for limited uptake of CAPS was the distance from young people’s homes to the CHIEDZA centre, the interruptions caused by COVID19 lockdowns and prevailing transport restrictions due to the pandemic.

This paper did not have a standard of care group and thus we could not compare outcomes of young people receiving community-based with those who received clinic-based ART. However, the viral suppression rates among those who were previously diagnosed and in care somewhere else were comparable with those receiving care at CHIEDZA. Also the small sample size and limited sociodemographic variables available did not allow a more in-depth analysis of risk factors for not linking to care.

## Conclusion

Our study provides evidence that provision of differentiated services for young people in the community is feasible across the whole HIV care cascade. In an era of widely available and free HIV testing young people diagnosed with HIV through a community based service are by definition harder to reach than those already diagnosed. Hence the linkage, retention and viral suppression outcomes achieved through this differentiated service delivery model among young people newly diagnosed with HIV need to be appraised in this context. Retention throughout the HIV-care journey for young people needs to become a priority programme goal in order to achieve the new UNAIDS 95-95-95 targets which are far from being reached among young people. Particular attention must be given to ensure young people newly diagnosed with HIV are linked to and retained in care.

## Data Availability

Individual, anonymised participant data and a data dictionary will be available through the London School of Hygiene & Tropical Medicine repository (Data Compass) at the time of publication.

## Acknowledgements

The Authors would like to acknowledge all the CHIEDZA clients, communities and staff for their contribution to this study.

## Authorship

RAF and RH conceptualised this study. RAF, MT, CMY, TB, CM and ED designed the study methodology. CDC and KK wrote the first draft of this manuscript. CDC, ED, OM and ES were responsible for oversight of the study implementation and administration with support form MT and CM. VS, TB and AP conducted data analysis. SB and CMY were responsible for the study process evaluation. FH, OM, LN, JM, RV and AC were responsible for the day to day activities of the study.

## Reference

1. Young people and HIV. Geneva: Joint United Nations Programme on HIV/AIDS (UNAIDS; 2021.

2. UNAIDS DATA. UNAIDS, Geneva, Switzerland; 2022.

3. 90-90-90 An ambitious treatment target to help end the AIDS epidemic. Geneva: Joint United Nations Programme on HIV/AIDS (UNAIDS). ; 2014.

4. 2025 AIDS Targets. UNAIDS, Geneva, Switzerland; 2020.

5. Summary sheet: Zimbabwe population-based HIV impact assessment ZIMPHIA 2020. 2020.

6. Summary sheet: Lesotho population-based HIV impact assessment LePHIA 2020. 2021.

7. Summary sheet: Malawi population-based HIV impact assessment MPHIA 2020-2021. 2022.

8. Summary sheet: Mozambique population-based HIV impact assessment INSIDA 2021. 2022.

9. Summary sheet: Uganda population-based HIV impact assessment UPHIA 2020-2021. 2022.

10. Chipanta D, Amo-Agyei S, Giovenco D, Estill J, Keiser O. Socioeconomic inequalities in the 90-90-90 target, among people living with HIV in 12 sub-Saharan African countries - Implications for achieving the 95-95-95 target - Analysis of population-based surveys. EClinicalMedicine. 2022;53:101652.

11. Boeke CE, Nabitaka V, Rowan A, Guerra K, Kabbale A, Asire B, et al. Assessing linkage to and retention in care among HIV patients in Uganda and identifying opportunities for health systems strengthening: a descriptive study. BMC Infect Dis. 2018;18(1):138.

12. Lamb MR, Fayorsey R, Nuwagaba-Biribonwoha H, Viola V, Mutabazi V, Alwar T, et al. High attrition before and after ART initiation among youth (15-24 years of age) enrolled in HIV care. Aids. 2014;28(4):559–68.

13. Desmonde S, Tanser F, Vreeman R, Takassi E, Edmonds A, Lumbiganon P, et al. Access to antiretroviral therapy in HIV-infected children aged 0-19 years in the International Epidemiology Databases to Evaluate AIDS (IeDEA) Global Cohort Consortium, 2004-2015: A prospective cohort study. PLoS Med. 2018;15(5):e1002565.

14. Low A, Teasdale C, Brown K, Barradas DT, Mugurungi O, Sachathep K, et al. Human Immunodeficiency Virus Infection in Adolescents and Mode of Transmission in Southern Africa: A Multinational Analysis of Population-Based Survey Data. Clin Infect Dis. 2021;73(4):594–604.

15. Leshargie CT, Demant D, Burrowes S, Frawley J. The proportion of loss to follow-up from antiretroviral therapy (ART) and its association with age among adolescents living with HIV in sub-Saharan Africa: A systematic review and meta-analysis. PLoS One. 2022;17(8):e0272906.

16. Haas AD, Radin E, Hakim AJ, Jahn A, Philip NM, Jonnalagadda S, et al. Prevalence of nonsuppressed viral load and associated factors among HIV-positive adults receiving antiretroviral therapy in Eswatini, Lesotho, Malawi, Zambia and Zimbabwe (2015 to 2017): results from population-based nationally representative surveys. J Int AIDS Soc. 2020;23(11):e25631.

17. Shanaube K, Macleod D, Chaila MJ, Mackworth-Young C, Hoddinott G, Schaap A, et al. HIV Care Cascade Among Adolescents in a “Test and Treat” Community-Based Intervention: HPTN 071 (PopART) for Youth Study. J Adolesc Health. 2021;68(4):719–27.

18. Shanaube K, Schaap A, Chaila MJ, Floyd S, Mackworth-Young C, Hoddinott G, et al. Community intervention improves knowledge of HIV status of adolescents in Zambia: findings from HPTN 071-PopART for youth study. Aids. 2017;31 Suppl 3(Suppl 3):S221–s32.

19. Mukora-Mutseyekwa F, Mundagowa PT, Kangwende RA, Murapa T, Tirivavi M, Mukuwapasi W, et al. Implementation of a campus-based and peer-delivered HIV self-testing intervention to improve the uptake of HIV testing services among university students in Zimbabwe: the SAYS initiative. BMC Health Serv Res. 2022;22(1):222.

20. McHugh G, Koris A, Simms V, Bandason T, Sigwadhi L, Ncube G, et al. On Campus HIV Self-Testing Distribution at Tertiary Level Colleges in Zimbabwe Increases Access to HIV Testing for Youth. J Adolesc Health. 2023;72(1):118–25.

21. Hensen B, Phiri M, Schaap A, Sigande L, Simwinga M, Floyd S, et al. Uptake of HIV Testing Services Through Novel Community-Based Sexual and Reproductive Health Services: An Analysis of the Pilot Implementation Phase of the Yathu Yathu Intervention for Adolescents and Young People Aged 15-24 in Lusaka, Zambia. AIDS Behav. 2022;26(1):172–82.

22. Chem ED, Ferry A, Seeley J, Weiss HA, Simms V. Health-related needs reported by adolescents living with HIV and receiving antiretroviral therapy in sub-Saharan Africa: a systematic literature review. J Int AIDS Soc. 2022;25(8):e25921.

23. MacCarthy S, Saya U, Samba C, Birungi J, Okoboi S, Linnemayr S. “How am I going to live?”: exploring barriers to ART adherence among adolescents and young adults living with HIV in Uganda. BMC Public Health. 2018;18(1):1158.

24. Casale M, Cluver L, Boyes M, Toska E, Gulaid L, Armstrong A, et al. Bullying and ART Nonadherence Among South African ALHIV: Effects, Risks, and Protective Factors. J Acquir Immune Defic Syndr. 2021;86(4):436–44.

25. Maskew M, Bor J, MacLeod W, Carmona S, Sherman GG, Fox MP. Adolescent HIV treatment in South Africa’s national HIV programme: a retrospective cohort study. Lancet HIV. 2019;6(11):e760–e8.

26. Enane LA, Davies MA, Leroy V, Edmonds A, Apondi E, Adedimeji A, et al. Traversing the cascade: urgent research priorities for implementing the ’treat all’ strategy for children and adolescents living with HIV in sub-Saharan Africa. J Virus Erad. 2018;4(Suppl 2):40–6.

27. Maskew M, Technau K, Davies MA, Vreeman R, Fox MP. Adolescent retention in HIV care within differentiated service-delivery models in sub-Saharan Africa. Lancet HIV. 2022;9(10):e726–e34.

28. Dziva Chikwari C, Dauya E, Bandason T, Tembo M, Mavodza C, Simms V, et al. The impact of community-based integrated HIV and sexual and reproductive health services for youth on population-level HIV viral load and sexually transmitted infections in Zimbabwe: protocol for the CHIEDZA cluster-randomised trial [version 1; peer review: 1 approved]. Wellcome Open Research. 2022;7(54).

29. Zimbabwe National Guidelines on HIV Testing and Counselling. Harare, Zimbabwe: Ministry of Health and Child Care; 2014.

30. Guidelines for Antiretroviral Therapy for the Prevention and Treatment of HIV in Zimbabwe. . National Medicines and Therapeutics Policy Advisory Committee (NMTPAC) and The AIDS and TB Directorate, Ministry of Health and Child Care, Zimbabwe; 2016.

31. Bochner AF, Meacham E, Mhungu N, Manyanga P, Petracca F, Muserere C, et al. The rollout of Community ART Refill Groups in Zimbabwe: a qualitative evaluation. J Int AIDS Soc. 2019;22(8):e25393.

32. Wilson KS, Mugo C, Moraa H, Onyango A, Nduati M, Inwani I, et al. Health provider training is associated with improved engagement in HIV care among adolescents and young adults in Kenya. Aids. 2019;33(9):1501–10.

33. Sabapathy K, Hensen B, Varsaneux O, Floyd S, Fidler S, Hayes R. The cascade of care following community-based detection of HIV in sub-Saharan Africa - A systematic review with 90-90-90 targets in sight. PLoS One. 2018;13(7):e0200737.

34. Barnabas RV, Szpiro AA, van Rooyen H, Asiimwe S, Pillay D, Ware NC, et al. Community-based antiretroviral therapy versus standard clinic-based services for HIV in South Africa and Uganda (DO ART): a randomised trial. Lancet Glob Health. 2020;8(10):e1305–e15.

35. MacPherson P, Lalloo DG, Webb EL, Maheswaran H, Choko AT, Makombe SD, et al. Effect of optional home initiation of HIV care following HIV self-testing on antiretroviral therapy initiation among adults in Malawi: a randomized clinical trial. Jama. 2014;312(4):372–9.

36. Labhardt ND, Ringera I, Lejone TI, Klimkait T, Muhairwe J, Amstutz A, et al. Effect of Offering Same-Day ART vs Usual Health Facility Referral During Home-Based HIV Testing on Linkage to Care and Viral Suppression Among Adults With HIV in Lesotho: The CASCADE Randomized Clinical Trial. Jama. 2018;319(11):1103–12.

37. Kopo M, Lejone TI, Tschumi N, Glass TR, Kao M, Brown JA, et al. Effectiveness of a peer educator-coordinated preference-based differentiated service delivery model on viral suppression among young people living with HIV in Lesotho: The PEBRA cluster-randomized trial. PLoS Med. 2023;20(1):e1004150.

